# A Renal Safety Checkpoint for Early High Intensity Statin Therapy in Critically Ill Patients With Acute Coronary Syndrome: A Multidatabase Target Trial Emulation

**DOI:** 10.64898/2026.08.05.26359828

**Authors:** Kaizong Huang, Xiaohan Zheng, Jie Liu, Chenyang Wu, Hong Sun

## Abstract

**Background:** High intensity statins are foundational after acute coronary syndrome (ACS), yet intensive care unit prescribing occurs while renal reserve, perfusion, and interacting therapies are changing. We tested a renal safety checkpoint integrating kidney status, hemodynamic instability, and drug interaction burden to identify when statin intensity may become nonexchangeable.

**Methods:** We emulated an active-comparator target trial across MIMIC-IV, eICU, and MIMIC-III. Critically ill adults with ACS, acute myocardial infarction, or percutaneous coronary intervention who received high- or moderate-intensity statins within 24 hours were included. The primary outcome was 7-day KDIGO stage 2 or 3 acute kidney injury or incident renal replacement therapy. Eligibility, time zero, treatment assignment, and follow-up were aligned. Database-specific propensity scores, overlap weighting, and standardization addressed confounding and treatment overlap. Safety domains, longitudinal analyses, bootstrap resampling, source omission, and endpoint sensitivities assessed robustness.

**Results:** Among 5,178 patients, 761 developed the primary outcome, including 223 who initiated renal replacement therapy. Standardized risks were 17.40% with high-intensity therapy and 15.01% with moderate-intensity therapy (risk difference, 2.39 percentage points [95% confidence interval (CI), -0.23 to 5.05]; risk ratio, 1.16 [95% CI, 0.99 to 1.39]). Risk separation was greatest with high hemodynamic instability (5.78 percentage points [95% CI, 1.56 to 9.74]) and high drug-interaction burden (6.24 percentage points [95% CI, -0.44 to 12.19]). Renal replacement therapy showed a 1.33-point risk difference (95% CI, 0.18 to 2.67).

**Conclusions:** This study moves statin safety assessment beyond fixed dose label or isolated creatinine measurement. The findings support a clinically actionable monitoring strategy in which early statin intensity is reassessed against evolving perfusion, kidney status, and interaction burden. This approach preserves intensive lipid lowering for physiologically suitable patients while identifying a high risk window in which temporary moderation.

**Clinical Perspective:** *What Is New?:*

- In critically ill patients with acute coronary syndrome, the average renal contrast between early high- and moderate-intensity statins was modest, while clinically important separation concentrated during hemodynamic instability and high drug-interaction burden.
- An active-comparator target trial framework linked statin intensity to 6-hour changes in perfusion, kidney function, and concomitant therapy, positioning renal tolerability as a context-sensitive treatment decision.

*What Are the Clinical Implications?:*

- High-intensity statin therapy remains the cardiovascular anchor, while a focused renal safety checkpoint can identify a transient window for closer surveillance and early intensity reassessment, followed by prompt intensification as physiology stabilizes.

## Introduction

Acute coronary syndrome (ACS) remains a major cause of death and recurrent cardiovascular events despite advances in reperfusion and secondary prevention. In a recent nationwide cohort, mortality at 1 year after acute myocardial infarction (AMI) remained 16% even among patients without cardiac arrest.^1^ Kidney injury adds an early and consequential risk. Among 455,806 contemporary percutaneous coronary interventions, acute kidney injury (AKI) occurred in 7.2%, and 0.7% required new dialysis.^2^ These hazards converge in intensive and coronary care, where ischemic protection begins during hemodynamic instability, contrast exposure, and evolving organ dysfunction. The treatment goal is to preserve cardiovascular benefit without compromising renal reserve.

Statins are central to this balance. They lower low density lipoprotein cholesterol and stabilize atherosclerotic plaque, with cardiovascular benefit emerging soon after ACS.^3,4^ Yet biologic exposure varies with dose, timing, duration, kidney function, circulatory state, and concurrent therapy. Ticagrelor can more than double rosuvastatin concentrations through transporter inhibition.^5^ Renal hypoperfusion, vasopressors, contrast, and nephrotoxins can further narrow treatment tolerance. The bedside question is not whether statins belong in ACS care. It is who can receive high intensity therapy immediately, when intensity merits reassessment, and how rapid lipid lowering can continue through transient organ vulnerability.

The 2023 European Society of Cardiology and 2025 American College of Cardiology and American Heart Association ACS guidelines recommend early high intensity or maximally tolerated statin therapy.^6,7^ They also emphasize lipid reassessment and further intensification when targets remain unmet. Yet acute renal safety thresholds and treatment choices during shock or evolving kidney injury remain less explicit. Randomized trials established intensive lipid lowering and flexible combination regimens.^8,9^ Procedural trials suggested that short course high dose statins may reduce contrast associated kidney injury.^10,11^ Randomized safety analyses support overall tolerability, whereas population data linked high potency initiation to early kidney injury admissions.^12,13^ Most studies used baseline exposure categories or selected procedural populations. Few captured changing treatment, time dependent confounding, and rapidly evolving physiology. Average effects may also conceal risk concentrated in patients with limited renal or circulatory reserve. The renal tolerability of high versus moderate intensity therapy during unstable ACS care remains unresolved.

We examined critically ill adults with ACS, AMI, or recent percutaneous coronary intervention (PCI) who received high or moderate intensity statin therapy within 24 hours. The primary outcome was severe AKI or incident renal replacement therapy (RRT) within 7 days. An active comparator target trial framework was coupled with sequential 6 hour assessment of treatment and evolving physiology. This approach focused on two accepted intensities while allowing perfusion, kidney function, and concomitant therapy to change. We tested whether statin intensity behaves as a physiology sensitive treatment decision and whether a renal safety checkpoint can preserve lipid lowering while identifying a transient window of reduced renal tolerance.

## methods

### Study Design and Target Trial Framework

We emulated a hypothetical active-comparator trial of early high-intensity versus moderate-intensity statin therapy. The assignment window extended from index time through 24 hours. Eligibility, treatment assignment, and follow-up were anchored to the same time zero. The primary estimands were the database-standardized 7-day risk difference and risk ratio for creatinine-based Kidney Disease: Improving Global Outcomes (KDIGO) stage 2 or 3 AKI or incident RRT. This estimand captured the renal consequence of selecting between two guideline-concordant strategies among patients with treatment equipoise.

### Data Sources

The analysis used harmonized, deidentified data from MIMIC-IV, the eICU Collaborative Research Database, and MIMIC-III. Each source contributed patient-level eligibility, treatment, covariate, and outcome information. Six-hour longitudinal records were linked when available. Source-specific treatment models retained local prescribing patterns, case mix, measurement practice, and renal event risk. Database standardization then translated the source estimates to a common target population.

### Eligibility, Index Time, and Cohort Construction

Eligible adults were admitted to an intensive care unit (ICU), coronary care unit, cardiac intensive care unit, or mixed intensive care unit during a qualifying ACS, AMI, or PCI hospitalization. Qualifying evidence comprised ST-segment-elevation myocardial infarction, non-ST-segment-elevation myocardial infarction, AMI with coronary angiography or PCI, PCI for ACS, or unstable angina with coronary investigation or treatment. Index time marked the earliest point at which all eligibility criteria were met. Treatment assignment began only after this point.

### Renal Risk Exclusions

Patients were alive and free of RRT at index time. Baseline serum creatinine was required from 24 hours before to 6 hours after index time. The primary cohort included patients with no AKI or KDIGO stage 1 AKI at baseline. Exclusions comprised KDIGO stage 2 or 3 AKI, active or chronic dialysis, end-stage kidney disease, kidney transplantation, recent major surgery, major trauma, burns, crush injury, rhabdomyolysis, severe active hepatic failure, and documented statin contraindications.

### Exposure Definition

The exposure window covered the first 24 hours after index time. High-intensity therapy comprised atorvastatin 40 to 80 mg daily or rosuvastatin 20 to 40 mg daily. Moderate-intensity therapy included atorvastatin 10 to 20 mg, rosuvastatin 5 to 10 mg, and guideline-recognized moderate-intensity doses of simvastatin, pravastatin, lovastatin, fluvastatin, and pitavastatin. Assignment followed the highest intensity recorded during the window. This strategy incorporated early escalation into the initial treatment decision.

### Outcome Definition

The primary renal outcome combined new creatinine-based KDIGO stage 2 or 3 AKI with incident RRT within 7 days. Incident RRT included continuous kidney replacement therapy, intermittent hemodialysis, or sustained low-efficiency dialysis. Baseline kidney function was established before treatment assignment. The harmonized endpoint used creatinine and RRT across all databases. Urine-output AKI was evaluated in sensitivity analysis.

### Covariates and Renal Safety Domains

Covariates represented demographic characteristics, baseline kidney function, chronic disease, acute severity, circulatory support, mechanical ventilation, vasopressor exposure, nephrotoxin burden, and selected missingness indicators. Three prespecified clinical domains integrated renal vulnerability, hemodynamic instability, and drug-interaction burden. Each domain was constructed before outcome estimation and classified into lower- and higher-risk states.

### Primary Analysis

Propensity scores for high-intensity therapy were estimated separately within each database. Overlap weighting emphasized patients who could plausibly receive either intensity. Standardized mean differences quantified covariate balance, and effective sample size summarized retained information. Weighted risks, risk differences, and risk ratios were estimated within each database. These estimates were standardized to the primary source distribution. Bootstrap resampling generated interval estimates for the primary contrast.

### Replication, Renal Safety Domains, and Longitudinal Analyses

Database-specific estimates were compared with an independent implementation of the standardized analysis. Fixed-effect and random-effects summaries quantified variation across databases. Leave-one-database-out analyses and a restriction to eICU hospitals with stronger representation of both strategies examined source influence. Renal safety contrasts were estimated within the three prespecified domains. A linked 6-hour module characterized the cumulative renal process and accompanying creatinine, RRT, mean arterial pressure, and vasopressor trajectories through 168 hours.

### Sensitivity Analyses

Prespecified analyses varied the renal endpoint, exposure classification, weighting approach, propensity-score specification, kidney function restriction, eligibility window, and clinical risk domain. RRT was examined as a severe standalone endpoint. E-values quantified the residual confounding strength required to explain the principal risk-ratio estimates.

## Results

### Cohort and Treatment Strategies

The study population comprised 5,178 critically ill adults with ACS, AMI, or PCI. eICU contributed 2,817 patients, MIMIC-IV contributed 1,780, and MIMIC-III contributed 581. High-intensity therapy was initiated in 4,025 patients and moderate-intensity therapy in 1,153. Both strategies were represented in each database, establishing an active clinical comparison across the three care settings (Figure 1a and 1b; Supplementary Tables S1 and S9a).

**Figure 1.**
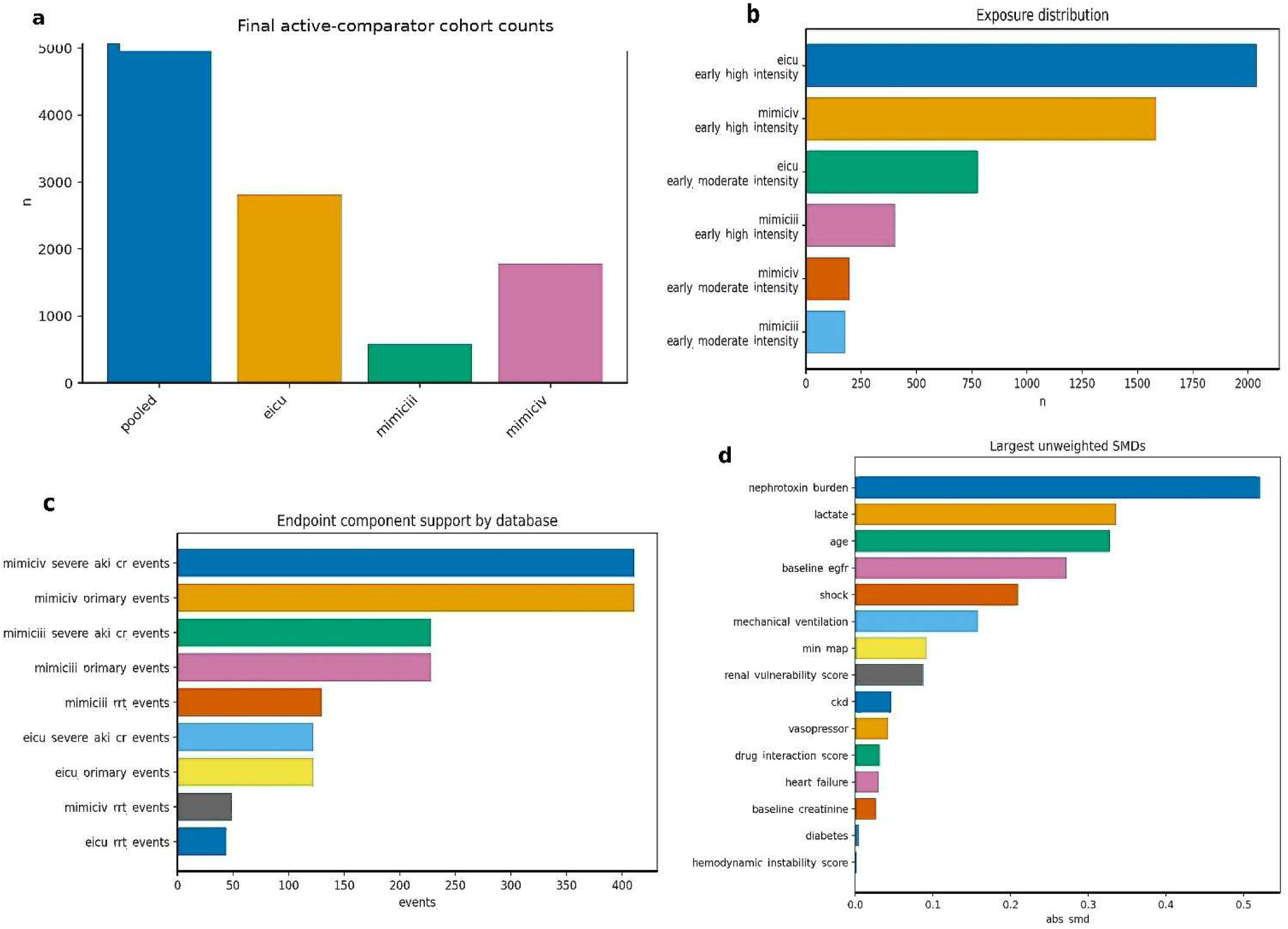
Cohort construction and analytic support. (a) Final ACS, AMI, and PCI ICU or coronary care unit cohort after harmonised eligibility criteria, baseline renal requirements, active comparator restriction, and outcome concordance. (b) High intensity and moderate intensity statin support by database during the first 24 hours. (c) Database specific support for the 7 day creatinine based severe AKI or RRT outcome and its components. (d) Baseline standardised mean differences before weighting. Together, the panels show the population, treatment contrast, renal outcome, and covariate structure used for estimation. AKI indicates acute kidney injury; Cr, Creatinine; RRT, renal replacement therapy; SMD, standardized mean difference; eGFR, estimated glomerular filtration rate; CKD, chronic kidney disease.

### Renal Outcome and Baseline Structure

The 7-day severe AKI or RRT outcome occurred in 761 patients, including 223 who initiated RRT. MIMIC-IV contributed 411 events, eICU 122, and MIMIC-III 228. Creatinine-based severe AKI or RRT accounted for every composite event. Baseline treatment differences were concentrated in nephrotoxin burden, lactate, age, and estimated glomerular filtration rate (Figure 1c and 1d; Supplementary Tables S2 and S9b).

### Primary Renal Estimate

Overlap weighting achieved excellent covariate balance across the three databases. The maximum absolute standardized mean difference was 0.007 overall and 0.0127 within any database. Standardized renal risk was 17.40% with high-intensity therapy and 15.01% with moderate-intensity therapy. The risk difference was 2.39 percentage points (bootstrap 95% confidence interval (CI), -0.23 to 5.05), and the risk ratio was 1.16 (95% CI, 0.99 to 1.39). The point estimate corresponded to approximately 24 additional severe renal events per 1000 patients treated with the high-intensity strategy (Figure 2a; Supplementary Figure S1 and S2; Supplementary Tables S3 and S10a).

**Figure 2.**
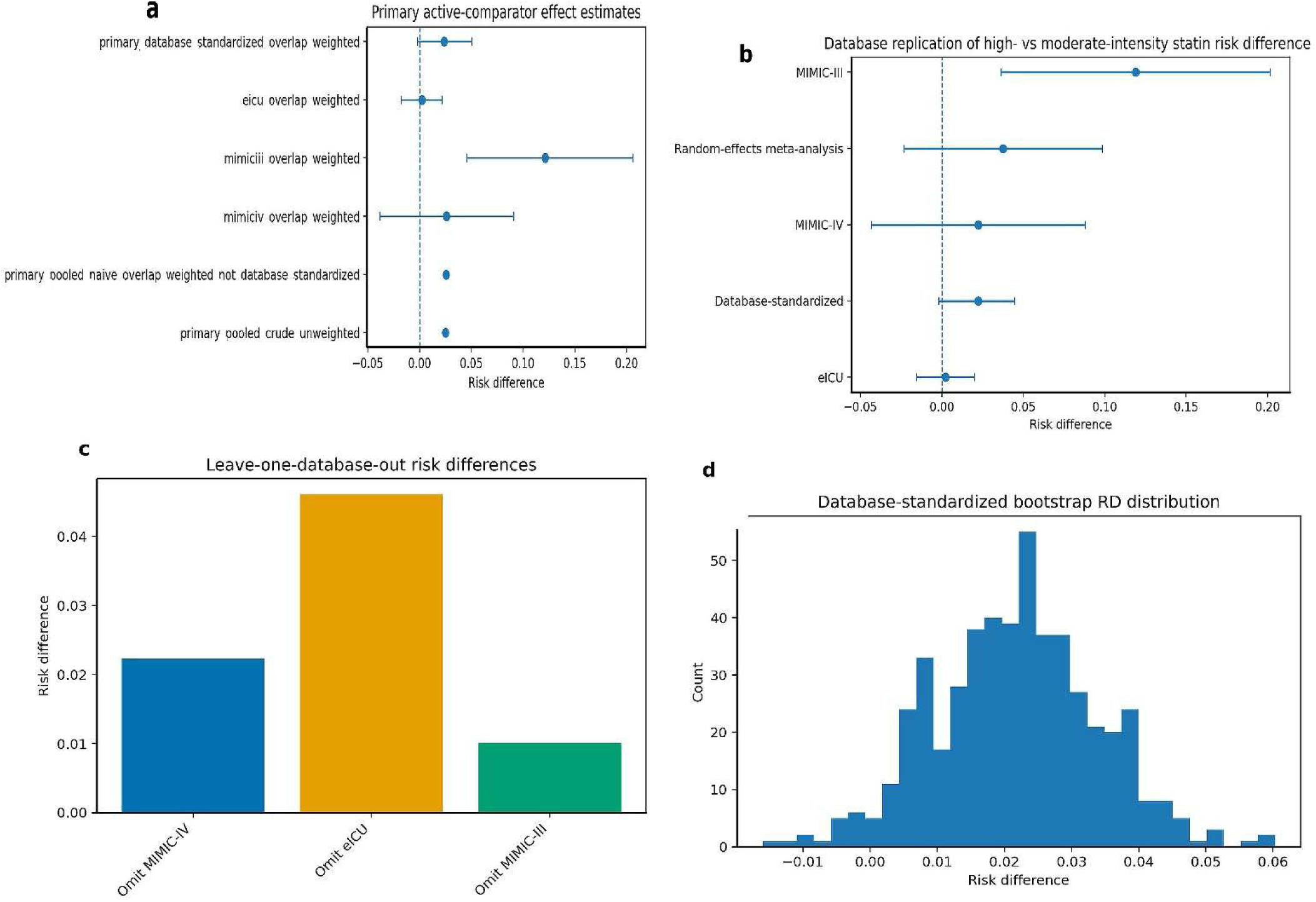
Primary estimate and multidatabase replication. (a) Prespecified database standardized overlap weighted estimates for high intensity versus moderate intensity statin therapy. (b) Source specific estimates and an independently implemented standardization analysis. (c) Leave one database out estimates characterizing source influence. (d) Bootstrap distribution of the standardized risk difference. The standardized estimand anchors the primary inference, while source specific results delineate heterogeneity and transportability.

### Across-Database Consistency

Estimates varied in magnitude across the three clinical environments. Risk differences were 0.25 percentage points in eICU, 2.60 points in MIMIC-IV, and 12.15 points in MIMIC-III. An independent implementation yielded a database-standardized difference of 2.23 points (95% CI, - 0.19 to 4.47). Leave-one-database-out estimates ranged from 1.00 to 4.61 points, and the bootstrap distribution remained centered near the primary estimate. Together, these analyses characterized the range of the treatment contrast across care systems while preserving the standardized population estimate (Figure 2b through 2d; Supplementary Figure S3; Supplementary Tables S6 and S10b).

### Renal Safety Pattern

The average contrast sharpened within prespecified clinical risk domains. Hemodynamic instability identified the clearest separation. The risk difference was -1.37 percentage points in the lower-instability stratum and 5.78 points in the higher-instability stratum (95% CI, 1.56 to 9.74). High drug-interaction burden showed a 6.24-point difference (95% CI, -0.44 to 12.19), compared with 1.69 points at lower burden. Renal vulnerability chiefly marked higher absolute event risk rather than a larger treatment contrast. This pattern linked early statin intensity to acute perfusion and medication context (Figure 3a; Supplementary Figures S3 and S4; Supplementary Tables S4 and S5).

**Figure 3.**
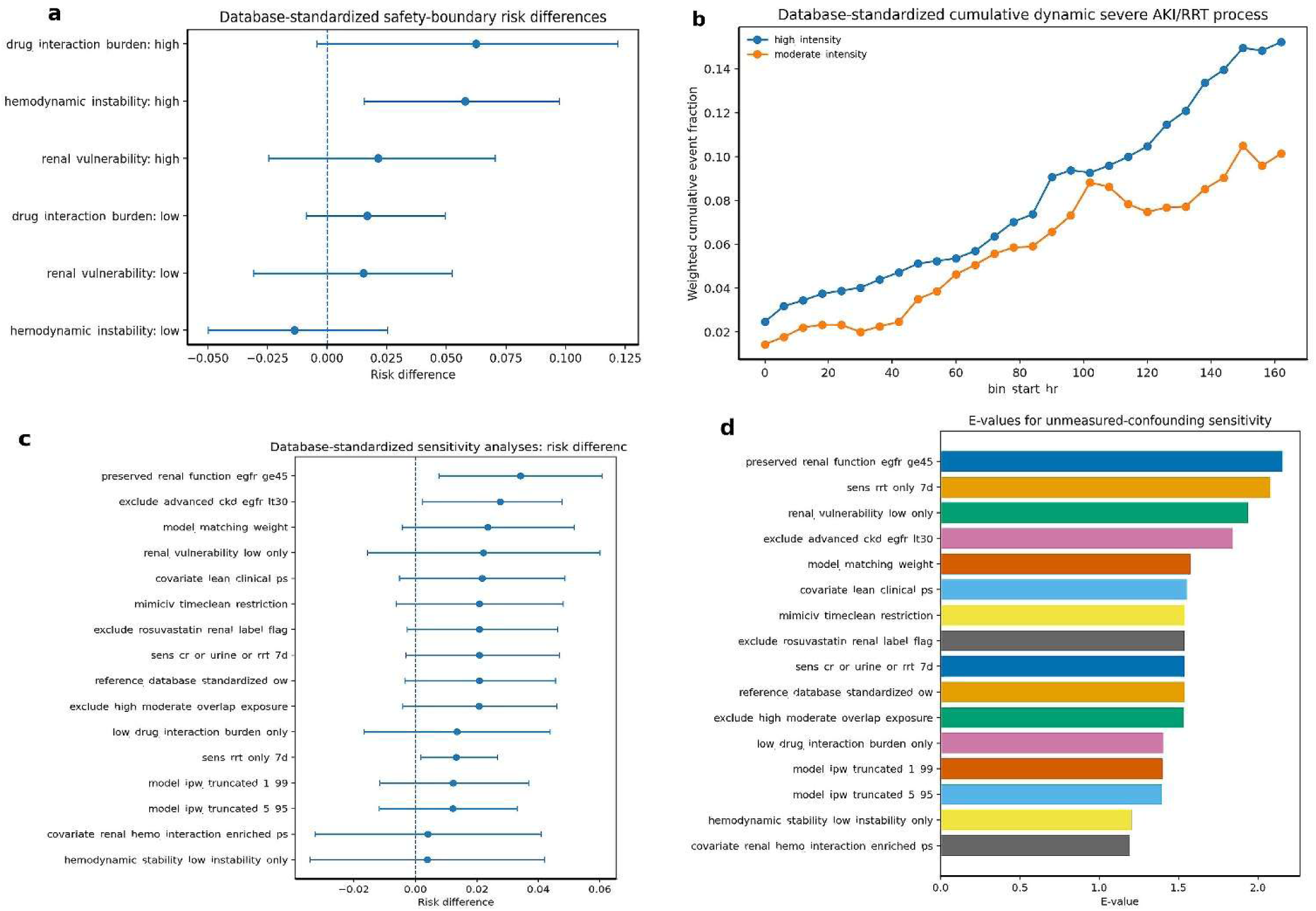
Context dependent renal safety pattern and robustness. (a) Prespecified estimates across renal vulnerability, hemodynamic instability, and drug interaction burden. (b) Database standardized cumulative severe AKI or RRT process across 6 hour follow up. (c) Prespecified sensitivity estimates. (d) E values for unmeasured confounding. The panels show where the standardized contrast becomes most apparent and how it behaves across time and analytic choices. AKI indicates acute kidney injury; RRT, renal replacement therapy; eGFR, estimated glomerular filtration rate; PS, Propensity score; Cr, Creatinine; IPW, inverse probability weighting.

### Longitudinal Renal Course

The longitudinal analysis included 52,678 patient-time rows from 5,177 cohort stays. The cumulative severe AKI or RRT process remained higher with high-intensity therapy throughout follow-up. Separation increased from 1.57 percentage points at 24 hours to 5.08 points at 162 hours. Time-updated creatinine, mean arterial pressure, vasopressor exposure, and RRT trajectories placed the widening contrast within the evolving critical care course. The trajectory paralleled the prespecified 7-day endpoint (Figure 3b; Supplementary Figure S5; Supplementary Table S7).

### Sensitivity Analyses

The renal pattern remained consistent across exposure, outcome, weighting, and kidney function analyses. RRT provided a concordant severe outcome, with a 1.33-point risk difference (95% CI, 0.18 to 2.67) and a risk ratio of 1.37 (95% CI, 1.04 to 1.98). Exposure exclusions, matching weights, and a lean clinical propensity-score model remained close to the reference contrast. As expected, the contrast narrowed after incorporating the prespecified clinical domains and among patients with lower hemodynamic instability. This alignment concentrated the renal signal within acute physiologic vulnerability. E-values were 1.53 for the reference estimate and 2.08 for RRT (Figure 3c and 3d; Supplementary Figure S6; Supplementary Table S8).

### Integrated Clinical Pattern

Across the complete evidence set, early high-intensity therapy showed modest average renal separation and a more pronounced difference during hemodynamic instability and high drug-interaction burden. The longitudinal course, RRT endpoint, and prespecified analytic variants converged on the same early clinical window. Renal safety therefore emerged as a context-sensitive intensity decision during the first days of unstable ACS care.

## Discussion

### Key Finding

This study identifies the early statin intensity decision in critical illness as a problem of clinical comparability, not a referendum on the cardiovascular value of intensive lipid lowering. Across 5,178 adults with ACS, AMI, or PCI, high intensity therapy was associated with a 2.39 percentage point higher 7 day risk of severe AKI or RRT. Separation was more pronounced during marked hemodynamic instability, reaching 5.78 percentage points, and the RRT analysis pointed in the same direction. The finding identifies an acute care boundary in which the renal tolerability of two guideline concordant intensities may diverge as perfusion reserve deteriorates.

The principal advance is a change in the clinical question. Earlier research largely asked whether intensive statin therapy is effective after ACS or whether statins are intrinsically nephrotoxic. Neither formulation captures the ICU decision, where statin treatment is already indicated and the relevant comparison is between two active, clinically acceptable intensities. By aligning eligibility, treatment assignment, and follow up at initiation, then examining severe renal events during the first 7 days, this study evaluates comparative tolerability when shock, contrast, vasopressors, nephrotoxins, and changing kidney function coexist. It shifts the evidence from a binary safety debate toward a context dependent treatment comparison.

This distinction helps reconcile prior renal evidence. PRATO ACS and TRACK D reported fewer contrast associated AKI events after short term high dose rosuvastatin in selected angiography populations,^10,11^ while patient level analyses from major ACS trials found no consistent excess kidney injury with higher potency therapy.^14^ Conversely, population data associated high potency statins with more AKI admissions, and rosuvastatin in sepsis associated acute respiratory distress syndrome raised renal and hepatic safety concerns.^12,15^ These studies addressed different purposes and physiologic states. Procedural prophylaxis and stable secondary prevention differ from critical illness, where perfusion failure, cumulative nephrotoxins, and drug interactions may dominate early renal risk. Our results connect these settings: the average contrast is limited, but becomes clinically visible when acute reserve is compromised.

The pattern across risk domains adds biological and clinical coherence. Hemodynamic instability was the clearest amplifier, supporting a perfusion dependent rather than fixed dose toxicity interpretation. Low flow and vasopressor dependence can intensify ischemic tubular stress, while contrast exposure and nephrotoxins add parallel injury. Reduced kidney function may narrow the exposure margin for high dose rosuvastatin, and transporter mediated interactions may increase systemic exposure; ticagrelor can increase rosuvastatin concentrations through relevant transport pathways.^16^ Renal vulnerability mainly identified higher background risk, whereas acute instability identified both higher risk and reduced comparability. Chronic kidney disease signals who is vulnerable; evolving physiology and medication context may signal when intensity merits reassessment.

### Clinical Implications

The findings support a renal safety adaptation strategy that preserves the cardiovascular priority of early statin treatment. With stable perfusion, preserved kidney reserve, and low interaction burden, early high intensity atorvastatin or rosuvastatin remains consistent with guideline based care. When shock physiology, evolving creatinine, heavy nephrotoxin exposure, severe renal impairment, or interacting medications cluster, immediate statin therapy may be paired with closer renal surveillance and an early reassessment of intensity. A temporary moderate intensity regimen may be considered in selected patients, followed by prompt escalation once perfusion, kidney function, and medication burden stabilize. This is not deintensification as a destination; it is sequencing of intensity across a rapidly changing risk window. RACING and LODESTAR support individualized lipid lowering architectures in stable disease.^8,9^ Our study extends that principle to early critical care.

Several design features strengthen this interpretation. Eligibility, time zero, exposure assignment, and follow up were aligned before estimation, reducing immortal time and treatment timing distortion.^17^ The active comparator avoided an artificial treatment versus no treatment contrast. Database specific propensity scores respected prescribing patterns and case mix, while overlap weighting centered inference on patients with treatment equipoise.^18,19^ Standardization produced a prespecified population average rather than a crude pooled estimate. Longitudinal trajectories assessed temporal coherence, and RRT provided a more severe outcome. Bootstrap, exposure, weighting, renal function, and source omission analyses tested robustness. These methods addressed a clinically relevant estimand: the 7 day renal consequence of initiating high versus moderate intensity therapy among patients for whom either strategy was plausible.

Variation across databases adds clinical depth to the standardized estimate. eICU contributed the largest and most geographically diverse population and moderated the average magnitude. MIMIC IV contributed a smaller positive estimate, while MIMIC III contributed stronger separation. Calendar era, case mix, contrast practice, creatinine measurement, RRT thresholds, and local prescribing patterns can influence effect magnitude across care systems. The standardized analysis preserved one common clinical question while retaining this real variation. Recurrent concentration of risk during hemodynamic instability carries greater clinical meaning than exact numerical agreement across sources. Longitudinal renal change and the RRT analysis reinforced this pattern. The result strengthens transportability. Readers can map the findings to local shock burden, renal reserve, concomitant therapy, and treatment practice.

### Limitations

This analysis combined an active comparator, aligned time zero, database specific propensity scores, overlap weighting, independent implementation, and extensive sensitivity analyses. Residual treatment selection may remain from procedural complexity, contrast dose, fluid strategy, shock severity, and clinician concern about kidney risk. Medication exposure and interaction burden were reconstructed from routine records, and the harmonized outcome prioritized creatinine based severe AKI and RRT. Domain and longitudinal analyses characterize clinical concentration and temporal coherence but do not establish biological mediation. Prospective contemporary ACS cohorts are needed to evaluate the proposed safety checkpoint.

## Conclusion

Across three critical care databases, early high intensity statin therapy showed a context dependent renal safety pattern, with the clearest risk separation during hemodynamic instability and high drug interaction burden. A proposed checkpoint integrating kidney status, perfusion, and interacting medications may preserve intensive lipid lowering while identifying patients who merit closer surveillance or temporary intensity moderation. The findings apply most directly to critically ill ACS, AMI, or PCI populations.

## Data Availability

The data analyzed in this study are available from PhysioNet to credentialed users who complete the required training and sign the applicable Data Use Agreement: MIMIC-IV, MIMIC-III, and the eICU Collaborative Research Database, where applicable. Patient-level derived data cannot be redistributed under the data-use terms.

## Supplemental Material

Supplementary Methods

Supplementary Results

Supplementary Figure S1-S6

Supplementary Table S1-S10

## Non-standard Abbreviations and Acronyms

ACS: acute coronary syndrome
AM: I acute myocardial infarction
AKI: acute kidney injury
PCI: percutaneous coronary intervention
RRT: renal replacement therapy
KDIGO: Kidney Disease: Improving Global Outcomes
ICU: intensive care unit
CI: confidence interval

